# A Critical Interpretive Synthesis to Develop Quality Assessment Tools for E-Cigarette Reviews: Scope and Protocol

**DOI:** 10.1101/2020.05.25.20112524

**Authors:** R. O’Leary, F. Costanzo

## Abstract

One component of a systematic review is the quality assessment of studies to determine their inclusion or exclusion. Studies on e-cigarettes are conducted in the contentious atmosphere surrounding tobacco harm reduction, which has resulted at times in research bias. Therefore, the quality assessment of studies on e-cigarettes requires more scrutiny than what is provided by generic tools on study design. This topic-specific quality assessment must examine the tests, measurements, and analysis methods used for their adherence to research standards. Furthermore, the studies need to be carefully screened for bias. Because standard quality assessment tools do not provide this topic-specific guidance, we propose to develop quality assessment tools specifically for reviews on e-cigarettes, and for our living systematic reviews on e-cigarettes for tobacco harm reduction.

## BACKGROUND

Tobacco smoking causes more than seven million deaths annually among the 1.1 billion smokers worldwide (World Health Organization, 2019a). The worldwide prevalence of cigarette smoking in 2018 was 27.5% for males aged 15 years old and over and 4.8% for females (World Health Organization, 2019b). Smoking has been demonstrated to be a causal factor in 38 diseases, with smoking the leading risk factor for cancers and chronic respiratory diseases, and the ninth leading factor for cardiovascular diseases worldwide (GBD 2015 Tobacco Collaborators, 2017). In 2017, the global burden of cancer deaths attributed to smoking was 63.17% of tracheal, bronchus, and lung cancers, and for deaths from respiratory diseases the prevalences were 38.6% of chronic obstructive pulmonary disease (COPD), 15.1% of tuberculosis, 11.86% of asthma, and 10.34% of lower respiratory infections (Institute for Health Metrics and Evaluation, 2020). Smoking diseases also cause suffering with years of life lived with disability (ibid).

Harm reduction is one of the strategies proposed for addressing the death and diseases caused by cigarette use. Tobacco harm reduction tactics include the substitution of lower-risk nicotine delivery products for cigarette smoking. Vapour products (e-cigarettes, here after termed “e-cigs”), invented in 2003 and widely marketed as a consumer product since 2010, are being investigated as a potential product for tobacco harm reduction (Abrams, Glasser, Pearson, et al., 2018; Farsalinos, 2018; Levy et al., 2018).

However, the use of e-cigs for harm reduction is a highly contested subject, often debated in a contentious and emotional atmosphere (Fairchild et al., 2018; Henningfield et al., 2018; Warner, 2019). In a survey of 81 health organizations’ position statements, only 30% supported e-cig use (Brady et al., 2019). “It appears that e-cigarettes will remain a controversial topic and heated debate will continue for many more years” (Farsalinos, 2018, p. 11).

Unfortunately, this atmosphere of contention is affecting research done in the area. Some researchers are buttressing their arguments against e-cigs with “strong assertions that go beyond the science (e.g., conflating correlation with causation, cherry picking results to highlight a particular viewpoint)…troubling trends that lead to greater confusion” (Abrams, Glasser, Villanti, et al., 2018, p. 90). The 2018 evidence review by Public Health England observed that

> the way the results of some studies with particular limitations have been designed and reported, and then subsequently presented in the media, may have caused serious concerns about EC use as shown by [the] growing *misperceptions* of the health risks of EC (McNeill et al., 2018, p. 172, emphasis added).

How the evidence on e-cigs is interpreted and (in)validated varies between researchers, leading to misperceptions and a lack of trust in the research (Brady et al., 2019). Therefore, assessing the available evidence on the health impacts of e-cig substitution for cigarette smokers entails more than simply gathering data, but also an examination of how the findings are reported.

We are joining the debate on tobacco harm reduction using e-cigs with a multi-topic project, *In Silico Science*, a series of systematic reviews on the respiratory, cardiovascular, and metabolic (weight) health effects of e-cigs substitution for cigarette smokers. These reviews are in development as a living systematic review, a review that is regularly updated with research studies as they are published, and the conclusions and recommendations revised as warranted by emerging data. All types of research designs with human subjects will be included: case studies, randomized controlled trials, clinical trials, observational studies, cross-sectional studies, user surveys, and qualitative studies. The contentious debate surrounding e-cigs means that the assessment of study quality is critical for the acceptance of our reviews as a source of trustworthy evidence and credible recommendations. To this end, we are developing a quality assessment tool for each review that will provide transparent and stringent criteria for the inclusion and exclusion of data. The first of these quality assessment tools will be for studies examining respiratory effects.

As for any systematic review, retrieving all the available studies is the first step. For our living systematic reviews, quality assessment determines which studies to include. Our reviews require a context-specific quality assessment tool that examines (1) the adherence to research design standards, (2) the appropriateness of tests and measurements, (3) the assessment of risk of bias in analyses, and (4) the assessment of risk of reporting bias in the findings and conclusions. This paper outlines the protocol which will be used to develop our quality assessment tool. This tool will be named REQUIRE (**RE**view of **QU**ality **I**n **R**esearch on **E**-cigarettes). REQUIRE will be developed with a critical interpretive synthesis literature review.

## METHODS

Critical interpretive synthesis is a literature review method created by Dixon-Woods et al. (2006). First and foremost, a critical interpretive synthesis is a review of concepts, not data, and quality assessment standards are concepts. A critical interpretive synthesis review can be conducted to develop a typology, and a quality assessment tool is both a classification and a theory of what constitutes sound research (Tricco et al., 2016). Another requirement for the review to develop REQUIRE is its ability to incorporate foundational concepts from different study types, and this functionality is one of the strengths of critical interpretive synthesis. Furthermore, the lines-of-argument synthesis technique of this method is well suited to assembling a topic-specific set of standards with inputs from the many available quality assessment and risk of bias tools (discussed below).

Also of importance, critical interpretive synthesis is iterative and reflexive. For the success of this review the review team must be willing to go back-and-forth between concepts and standards and to spend considerable time in discussion. The complexity inherent in synthesizing the components of REQUIRE will necessitate an extended dialogic process (Plamondon et al., 2019).

Critical interpretive synthesis has been employed previously for the creation of standards and guidelines in health research and healthcare (Ako-Arrey et al., 2016; Catalano et al., 2018; Lippmann et al., 2019; Morgan et al., 2018; Mothupi et al., 2018; Plamondon et al., 2019). Considering the fitness-for-purpose of critical interpretive synthesis, we have chosen it to develop REQUIRE.

In addition to the methodology of critical interpretive synthesis, the development of our quality assessment tool is informed by Whiting et al. (2017) “A proposed framework for developing quality assessment tools.” This framework provides pragmatic steps for the process of developing the tool, while critical interpretive synthesis supplies the methods for the search strategies, the development of inclusion criteria, the synthesis of the quality assessment components, and the development of the quality assessment rating scales.

This protocol identifies the processes to develop REQUIRE-R, the protocol for the respiratory review. The processes for developing REQUIRE-C (cardiovascular effects) and REQUIRE-M (metabolic/weight effects) will follow the same course except for the reviews selected for the creation of the inventory library (Process Two) and the topic-specific search items for the research standards for tests measurements, and analysis methods (Process Three).

## REVIEW PROCESSES

There are seven planned processes for the development and utilization of the quality assessment tool. They are

1. assembling published quality assessment tools
2. creating an inventory of study designs, tests, measurements, and analytical techniques
3. identifying research standards for the inventory items
4. formulating risk of bias indicators
5. designing the tool through synthesis
6. pilot testing and revision
7. knowledge translation

<u>Process One</u> – retrieving published tools for research design quality assessment and risk of bias. The iterative search strategies of critical interpretive synthesis will be a key component of this phase of the review.

Quality assessment tools will be collected by the research leader who has expertise in this area. Many quality assessment tools are well established, some published as collections including *CASP Appraisal Checklists* (Critical Appraisal Skills Programme, 2018), *NICE Process and Methods Guides* (National Institute for Health Care and Excellence, 2015), *CLARITY Group Instruments* (Evidence Partners, 2017), and the *JBI Critical Appraisal Tools* (Joanna Briggs Institute, 2017). Other tools are available for individual research designs, such as *ROBINS-I* (Sterne et al., 2016) and the recently developed *MASTER* (MethodologicAl STandard for Epidemiological Research, Stone et al., 2019). A bibliography of guidelines is available from the EQUATOR Network (https://www.equator-network.org/).

Risk of bias indicators will be a key component of our tool. Several checklists and references are available to assess risk of bias, notably the widely used *Cochrane RoB2* (Cochrane Training, 2019). The Centre for Evidence Based Medicine *Catalogue of Bias* (2020) provides full descriptions and examples of 58 different biases. Risk of bias assessments in REQUIRE will be applied to examine the validity of study analyses and identify bias in the reporting of analyses and conclusions.

<u>Process Two</u> – developing an inventory of research designs, tests and measurements, and analysis methods. This three-step process will develop a sampling frame of studies, perform a data extraction of the items, and compile the items identified into the inventory.

*Create the sampling frame:* The production of the inventory will be drawn from a sampling frame of studies produced in two steps. First, a library of studies meeting the inclusion criteria (see below) will be retrieved from the four most recently published reviews that address e-cigs and respiratory effects and in addition represent differing positions on tobacco harm reduction: Bals et al. 2019 (European Respiratory Society, cautionary, risk-adverse position); Gotts et al. 2019 (nicotine abstinence position); National Academies of Sciences, Engineering, and Medicine 2018 (risk evidence statements); and Polosa et al. 2019 (pro tobacco harm reduction position).

The title and abstract inclusion criteria are (1) research specifically on e-cigs, (2) human-based research, (3) reporting data on any respiratory health effect or respiratory test, and (4) published starting from 2016. The study publication date criterion excludes earlier research based upon older models of e-cigs which are no longer on the market (Strongin, 2019; Williams & Talbot, 2019). Earlier research, for the most part, is not applicable because different models generate aerosols with different characteristics (Wagener et al., 2017). Only newer studies are likely to have research based upon e-cigs currently used by smokers. Therefore, studies conducted prior to 2016 and research done on first- and second-generation e-cigs will be excluded. Also excluded are studies on EVALI, a US disease outbreak associated with vaping, because the disease is attributed almost exclusively to the consumption of illegal THC products, not commercially available e-cigs (US Centers for Disease Control and Prevention, 2020). The studies meeting the inclusion criteria will be entered into Mendeley bibliographic software.

### Generate items

The studies in the library will be used as a sampling frame to provide an inventory of research designs, tests, measurements, and analytical techniques. A data extraction form will be developed and pilot tested on five studies, and revised as needed. Some studies may be excluded at this step because they lack data on health effects.

### Assemble inventory

The inventory will be compiled from all the items identified in the data extraction of studies in the sampling frame. Some items may be excluded if they appear to be a one-off measurement or test.

<u>Process Three</u> – A search for research standards for tests, measurements, and analytical methods. The quality assessment tools retrieved in the first process step are general; REQUIRE must have assessments of the specific tests and measurements in the studies of respiratory health impacts. Each team member will conduct searches focused on research standards for the inventory items through multiple strategies:

* conducting keyword searches of item names in Google Scholar and PubMed
* reference checking and citation chasing of journal articles in the inventory library
* collecting comments on study quality in the reviews used for the sampling frame
* hand-searching of selected journals (see Appendix 1)
* scanning respiratory textbooks (see Appendix 2)
* reviewing government and industry publications on e-cigarette product standards
* obtaining recommendations from experts in respiratory health.

In consideration of the need for reflexivity in the standards search, the research team will meet regularly to discuss search strategies and share other potential sources of information.

The search for quality tools and research standards will continue until saturation appears to have been achieved as indicated by diminishing retrievals, or until the project timeline of four weeks for this process has been reached. The retrievals will be deduplicated and collated.

<u>Process Four</u> – Formulating risk of bias indicators. The formulation of explicit indicators of biases in analysis and reporting will be based on the tools identified in the first process step and observations made by the researchers from their reading of the studies.

The biases of researchers about tobacco harm reduction may intentionally or unintentionally result in faulty analyses with “data cooking” by excluding some data or “data beautification” by overstating the significance of statistical tests (Chevassus-au-Louis, 2019). Researchers may also introduce reporting bias by specifying particular outcomes for clinical trials (Richards and Onakpoya, 2019), thereby excluding analyses of data not conforming to their position on tobacco harm reduction. Due to researchers’ highly emotional stance on tobacco harm reduction (Warner, 2019), spin bias, a type of reporting bias, may occur through “the intentional or unintentional distorted interpretation of research results, unjustifiably suggesting favourable or unfavourable findings that can result in misleading conclusions” (Mahtani et al., 2019, para. 1). This is the reporting bias discussed above in the Background section.

The critical interpretive synthesis method of reflexivity will be central to the process of formulating indicators of bias. After reviewing the tools for the risks of bias, each researcher will draft a list of indicators of bias in analyses, and a separate list for reporting biases based on these tools and their observations from reading the studies. These lists will be compared and discussed until consensus is reached on the bias indicators. To address the potential bias within the team due to its pro-harm reduction position, reporting bias indicators will be worded generically to be equally applicable to both pro and anti e-cig positions. Our reflexivity must include questioning our own positions (Kozlowski, 2016).

<u>Process five</u> – construction of REQUIRE. This process will have two steps: the first is the synthesis of quality assessment tools, research standards, and bias indicators, and the second is the development of a quality rating scale.

### Tool synthesis

REQUIRE will be synthesized from items selected from the inventory of quality assessment tools, the research standards, and the indicators of bias. The research team will make judgements with constant comparative analysis conducting a lines-of-argument synthesis on which components of the quality assessment tools will be incorporated and which research standards appear to be the most widely accepted.

These items will be assembled into two sections. One will contain the quality assessment standards for each study design and the bias indicators. The number of items in this section will be closely monitored to minimize evaluator burden. The second section will be a list of the research standards for the tests, measurements, and analysis techniques specific to the topic area. Clarity in the descriptions of the items will be paramount to insure the reliability of the tool.

The synthesis process will entail multiple rounds of discussion among the research team and scheduled periods of time for the researchers to reflect on the selection of items. These two activities will facilitate the reflexivity central to the critical interpretive synthesis review. Once the synthesis is complete, a study quality assessment form will be formatted with the relevant items in four domains: research design, tests and measurements, analysis, and reporting. The research team will review the forms for their completeness and fitness for purpose.

### Rating scale

Once the items in REQUIRE have been synthesized and formatted, then there is the critical step of formulating a rating scale. A numerical rating system is not under consideration because these scales have been shown to be unreliable (Stone et al., 2019; Whiting et al., 2017). The preliminary plan for the rating scale is standards-based and specified for each domain. No overall study quality assessment will be made.

The ratings scale will be formulated from the study’s adherence to the quality assessment items as developed in the synthesis. The research design rating will be based on the fulfillment of required criteria. The tests and measurements will be individually rated based on the identified research standards. The risk of bias assessments for the two domains of analysis and reporting of findings and conclusions will be based on the absence or presence of bias indicators. The preliminary quality rating scale for each domain and the actions to be taken based on the quality assessment for our living systematic review are presented in Table 1 below.

**Table 1.** – Quality Assessment Ratings

| Rating | Action for Living Systematic Review |
| --- | --- |
| **Research design** |  |
| Not meeting standards | Excluded from review, studies listed in appendix |
| Meeting standards | Included in review |
| Robust standards | Highlighted in review |
| **Tests and measurements** |  |
| Not valid | Data excluded from analysis, studies listed in appendix detailing the excluded tests |
| Weak | Data analyzed separately |
| Valid | Data included in analysis |
| Strong | Data included in analysis and a sub-group analysis |
| Not assessed | Data referenced in discussion |
| **Analysis** |  |
| Low risk of bias | Findings included in narrative analysis |
| Bias concern flagged | Findings referenced in discussion |
| High risk of bias (multiple bias indicators) | Analysis excluded, studies listed in appendix |
| **Reporting Analyses and Conclusions** |  |
| Low risk of bias | Included in narrative analysis |
| Bias concern flagged | Referenced in discussion |
| High risk of bias (multiple bias indicators) | Identified in discussion as invalid. |

As indicated by the quality assessment ratings, some studies may be excluded in their entirety. Some included studies may provide only valid tests, while other included studies may provide valid tests and analysis but have their findings and/or conclusions bracketed or excluded.

To reiterate, these are preliminary categories and terms for the quality assessment scales. They may be changed or further developed during the synthesis process and/or during the pilot testing process discussed next.

<u>Process Six</u> – pilot testing. REQUIRE will be pilot tested by the research team and then circulated for internal peer review.

### Review Team Pilot Testing

The pilot test will have two steps. For the first step, two research team members will test the quality assessment form for each research design on three studies chosen at random from the Inventory Library. The researchers will report the time taken to complete the assessments, compare the inter-rater reliability, and record observations on its use. The full team will review the pilot testing reports and revise the assessment items and form as needed. In the second step, the revised form will be retested by each research team member on one study for each research design, and again revised as needed after discussion.

### Internal peer review

REQUIRE and its quality assessment form will be distributed to a minimum of two internal reviewers for feedback. Depending on the extent of the feedback, the tool will be revised, or if substantive changes are recommended, then the second step of pilot testing will be conducted, and the revisions circulated to the internal peer reviewers for approval.

<u>Process Seven</u>: utilization and knowledge translation. This process will be conducted both internally and with publications and networking.

The first knowledge translation action will be the registration of the protocol in PROSPERO. The protocol will then be submitted to an open access journal. We also anticipate publishing an article on the tool development process.

For our purpose, REQUIRE-R will be utilized for our living systematic review on the effects of e-cig use on respiratory health. The reviewers will be tasked with reporting on the reliability and evaluator burden of using the tool, and it may be further revised. For the planned living systematic reviews on cardiovascular effects and metabolic (weight) effects, REQUIRE will be reworked to incorporate the tests and measurements for those topic areas by conducting the steps in Process Two to assemble a sampling frame and Process Three to identify research standards on the subject-specific items.

After REQUIRE has been evaluated in our first living systematic review of the series, we will publish it on a dedicated website as REQUIRE-R (respiratory effects). To publicize the tool and the website, we will send an e-mail blast to the corresponding authors of the studies in the Inventory Library. We will publish REQUIRE-R in a journal (to be selected). We will inquire with EQUATOR on the possibility of having it linked on their website.

These activities will be repeated for REQUIRE-C (cardiovascular effects) and REQUIRE-W (metabolic/weight effects). On the website we will solicit feedback on the tool and will consider revising it before the end of our project (August 2022). Plans for archiving REQUIRE will need to be developed so that it remains accessible after our project has concluded.

For future research, REQUIRE could be expanded to assess the study quality of human-subject research beyond e-cig research. The development of a quality assessment tool for studies of respiratory or cardiovascular health would facilitate reviews conducted in those areas.

## CLOSING COMMENTS

E-cig research is a controversial field, and individual studies are being published in support of and against the use of e-cigs in tobacco harm reduction. A living systematic review can inform the debate. Yet a systematic review of available studies will fail to provide credible evidence unless it weeds out research with substandard study designs, weak testing methods, biased analyses, and skewed reporting. REQUIRE, a review of the quality in research on e-cigarettes, will serve as a standard to assess published studies. The quality assessment provided by REQUIRE will allow for the inclusion of robust data even where reporting bias has coloured the findings. For our living systematic reviews, retrieving all the available studies with their data is only the first step - identifying sound data with REQUIRE will be key to gaining trust in the quality of the findings and conclusions of our reviews.

### FUNDING SOURCE

The protocol was produced with the help of a grant from the Foundation for a Smoke Free World, Inc. The contents, selection and presentation of facts, as well as any opinions expressed in the protocol are the sole responsibility of the authors and under no circumstances shall be regarded as reflecting the positions of the Foundation for a Smoke-Free World, Inc. The Grantor had no role in the selection of the research topic, study design, or the writing of the protocol or the living systematic review project.

### CONFLICT OF INTEREST

The authors declare no conflict of interest.

## Data Availability

protocol

## Acknowledgement

The authors wish to than Gianluca Conte for contributing Appendix 2 and Sandra Marquis, PhD for her editing support and comments.

## Appendix 1: Hand Search Journals

| Journal title | Web address |
| --- | --- |
| **Advances in Respiratory Medicine** | https://journals.viamedica.pl/advances_in_respiratory_medicine |
| **European Respiratory Journal** | https://erj.ersjournals.com/ |
| **Expert Review of Respiratory Medicine** | https://www.tandfonline.com/loi/ierx20 |
| **International Journal of COPD** | https://www.dovepress.com/international-journal-of-chronic-obstructive-pulmonary-disease-journal |
| **Pulmonology** | https://www.sciencedirect.com/journal/pulmonology |
| **Respiratory Research** | https://respiratory-research.biomedcentral.com/ |
| **Respirology** | https://onlinelibrary.wiley.com/journal/14401843 |
| **The Lancet Respiratory Medicine** | https://www.thelancet.com/journals/lanres/onlineFirst |

## Appendix 2 – Respiratory Textbooks

## Notes

### Competing Interest Statement

The authors have declared no competing interest.

### Author Declarations

exempt protocol - no human subjects

